# What works, for whom, and under what circumstances for recipients of training in opportunistic behaviour change conversations: a mixed methods realist evaluation protocol

**DOI:** 10.64898/2026.03.19.26348867

**Authors:** Beth Nichol, Angela M. Rodrigues, Robert Anderson-Weaver, Sonia Dalkin, Rebecca Hunter, Heather Brown, Christian Morgner, Beth Stuart, Charlotte Albury, Catherine Haighton

## Abstract

**Background:** ‘Making Every Contact Count’ (MECC) is a person-centred initiative that enables service providers across settings to support behaviour change through conversations about health and wellbeing. MECC has been widely implemented across the UK and internationally, although training approaches vary considerably and do not consistently translate into MECC delivery. Evidence suggests that Healthy Conversation Skills (HCS) training, which supports service users to identify their own solutions, may be an acceptable and effective means of delivering MECC across settings. This realist evaluation aims to understand which elements of HCS training work, for whom, under what circumstances, in what respects, to what extent, and why, to inform the adaptation of HCS across settings to ensure that all recipients are equipped to deliver MECC.

**Methods:** This mixed-methods realist evaluation will comprise pre- and post-training surveys (at baseline, immediately post-training, and approximately eight weeks post-training) and realist interviews. Two participant groups were selected for comparison: service providers working or volunteering in the voluntary, community, and social enterprise (VCSE) sector, and undergraduate pharmacy students. Initial programme theories were developed through abductive reasoning, literature scoping, and stakeholder engagement. Survey data will assess outcomes of HCS training, while realist interviews will explore how these outcomes are generated by underlying mechanisms within specific contexts.

**Discussion:** A refined programme theory will be produced, explaining how and why HCS training leads to MECC delivery across different settings. Findings will inform how HCS training can be adapted for distinct audiences, identify the core components of MECC training that must be preserved, and guide future evaluations by examining whether HCS training translates into sustained MECC delivery. The findings of this study will inform resource allocation for preventative health interventions outside of healthcare settings and thus have the potential to shape public health policy, empower non-specialist providers, and strengthen strategies for disease prevention.

## Introduction

The World Health Organisation has described interventions that target smoking, alcohol intake, physical activity, and diet as amongst the ‘best-buys’ in reducing the rising incidence of non-communicable diseases (NCDs) and their associated costs (1). Whilst upstream interventions that address the structural drivers of the rise of NCDs are key to addressing them (e.g. political, economic, or policy changes such as smoke-free legislation) (2), progression of upstream interventions is slow and faces several barriers including conflicting political interests with evidence, and opposing public opinion (2). Downstream interventions (including individual level interventions) can support people to make changes to their behaviour and create a culture change towards prevention rather than treatment of diseases (3, 4). Concurrently, in the United Kingdom (UK) the most recent 10-Year Health Plan continues to promote person-centred care (placing individuals’ needs and wants at the forefront) and autonomy (encouraging individuals to be actively involved in their health and wellbeing) (3).

Making Every Contact Count (MECC) is a person-centred intervention that aims to utilise existing conversations to promote health behaviour change (5). Its alignment with strategic focuses of prevention and autonomy mean that MECC continues to be implemented across the UK and beyond, since it became compulsory in 2016 that all NHS staff are trained to deliver MECC to their patients (5). A unique feature of MECC is that it does not require its deliverers to be specialists in behaviour change or healthcare professionals. It’s approach, resembling motivational interviewing through asking open questions as opposed to providing advice, means that in theory anyone can deliver and receive MECC (6). Subsequently, MECC has been implemented across a range of settings outside of healthcare, including fire and rescue, local authorities, and the voluntary, community, and social enterprise (VCSE) sector (7, 8). However, it is noteworthy that there is a lack of evidence to support sustained behaviour change in recipients of MECC. To date, there has only been one fully powered controlled trial which was not randomised (9). There remains a need for a fully powered trial that examines the effectiveness of MECC for the general population, to determine optimal allocation of health and social care funding to reduce NCDs. However, given the varied approaches to MECC training, understanding training mechanisms is foundational to ensure that MECC training facilitates delivery of MECC across settings, before investing in costly effectiveness trials.

A frequent challenge with implementing MECC is the barriers to service providers delivering MECC to patients following MECC training (10, 11). Frequently reported barriers to delivery of MECC across settings include lack of time (10, 12–16), a perception that MECC is not part of the service providers’ role (10, 13, 14, 17), and lack of confidence (7, 10, 13). Compounding this challenge, MECC has until recently been poorly defined (6), and subsequently the content of MECC training varies widely across regions, with many regions developing their own training programme (13). In accordance with national guidance for MECC training (18), many MECC training programmes focus on providing information to enable service providers to provide brief advice and signposting (4, 19, 20). By contrast, Healthy Conversation Skills (HCS) is a specific approach to MECC training that reflects motivational interviewing skills (21). Thus, it complements the recently agreed definition of MECC amongst experts (6). It is also the most appropriate and empirically supported available training, having been repeatedly demonstrated to increase confidence (22, 23) and competence (21, 22, 24–26) of service providers to deliver MECC, encouraging MECC delivery (25). HCS is also the most feasible approach to deliver MECC at scale, as its avoidance of advice provision makes it appropriate for service providers who are not healthcare professionals and therefore do not feel confident enough or willing to provide advice.

However, the broad application of HCS across settings requires nuanced investigation, as it is conceivable that HCS training impacts different audiences in different ways. For example, service providers from the VCSE may require experiential learning, whereas healthcare professionals may be more comfortable with a ‘see one, do one’ approach (27). Equally, service providers from the VCSE often feel confident in signposting but may feel unsure about proactively facilitating goal setting (28), whereas healthcare professionals may be familiar with goal setting but find it difficult to avoid providing advice (29). Also, initially delivered across three separate sessions of three hours each, HCS has also been shortened to one three-hour session (HCS ‘lite’), although this shortened iteration has yet to be evaluated. Whilst this facilitates the delivery of HCS training at scale, as multiple sessions may be simply unfeasible for many VCSE organisations, it is uncertain whether it is sufficient to encourage recipients to deliver MECC. A recent review, collating all studies examining the effect of MECC training on the confidence and competence of trainees (20), did little to explain what elements worked for whom, in what circumstances, and why, which is particularly concerning given the aforementioned diversity in approaches to MECC training.

Thus, to address this explanatory gap about the mechanisms of HCS training within different contexts, the current study adopts a realist evaluation approach to understand ‘*what* [elements of HCS training] *works, for whom, under what circumstances, in what respects, to what extent, and why?*’ (30). This evaluation will compare service providers from the VCSE, who are often highly experienced in interacting with service users but may not be confident in health promotion, with undergraduate pharmacy students, who have a healthcare background but may have little experience of interacting with service users. Pharmacy students have been selected as the healthcare professional comparison group as there is no available MECC literature that focuses on pharmacists despite implementation of MECC within this area (5, 20). This realist evaluation will test and refine programme theory explaining how and why HCS training enables MECC delivery. The resulting programme theory will inform on how HCS training can be adapted for different audiences, to make sure that it equips all recipients with the knowledge, confidence, and motivation to deliver MECC. In addition, the findings will also facilitate the future evaluation of MECC delivery in the VCSE, by exploring whether HCS training is sufficient for MECC delivery. Practically, this evaluation will also inform to what extent HCS training can be adapted (or shortened) without losing its impact. The findings will also provide utility for other approaches to MECC training, by identifying important elements needed to equip staff to deliver MECC.

## Methods

A realist evaluation approach has been selected as appropriate for the current study. Namely, the realist evaluation will test and refine initial programme theories (IPTs) using context, mechanism, outcome (CMO) configurations (31), which have been developed through stakeholder involvement and scoping of the existing literature. These CMOs describe in-depth, in written format, the specific context that is required for a specific part of the HCS training (mechanism resource) to create a specific response (mechanism reasoning) that results in the desired outcome. Importantly, individual CMO configurations will also be presented visually through an overall programme theory for each participant group. RAMESES II guidance (32) was adhered to for reporting of this protocol, and the checklist will be completed on publication of the final publication of the evaluation and its findings. Prior to data collection for interviews and before accessing the quantitative data, the evaluation was pre-registered on Open Science Framework (OSF): https://osf.io/wpbn3/overview. Recruitment and data collection began in August 2025 and is ongoing, due to be completed by July 2026. Results are expected by March 2027.

### Study design

This study is a concurrent mixed methods realist evaluation, consisting of two methods of data collection: 1) a quantitative before and after survey, and 2) qualitative interviews with trainees. Both methods of data collection will sample VCSE service providers and pharmacy students evenly, for comparison. The quantitative survey will be used to generate data on the extent and distribution of outcomes and how these differ between VCSE service providers and pharmacy students (context) (33), whilst the realist interviews will help to explain how and why these outcomes are produced, by linking them to CMO configurations. The quantitative survey was designed to be complimentary to existing evaluations of HCS training, to also aid comparison between groups of trainees (23, 26, 29). A between-subjects design for the quantitative survey will aid comparison between participant groups. Ethical approval has been obtained from Northumbria University prior to recruitment and data collection (Project ID: 10429).

### Study setting

Data collection will utilise scheduled (for the quantitative surveys) and previously delivered (for the interviews) HCS ‘lite’ training across Portsmouth, delivered by Portsmouth City Council. Portsmouth covers a diverse area in the South of England, although is currently in the early stages of implementation to the VCSE. Additionally, training will be delivered to pharmacy students at Newcastle University, which is situated in the North East of England and is surrounded by rural and coastal communities. Both settings were selected for pragmatic reasons; utilising existing delivery of HCS ‘lite’ by RAW across Portsmouth, and opportunities to deliver HCS ‘lite’ in Newcastle, where the lead author (BN) is based.

### Participants

All participants must be over 18 and have attended (or about to attend) HCS ‘lite’ training in Portsmouth, delivered by RAW. Additionally, participants must either be a pharmacy student or work or volunteer within the VCSE at the time of receiving HCS ‘lite’ training.

### Healthy Conversation Skills ‘lite’ training

This evaluation utilises existing scheduled HCS ‘lite’ training (a shortened version of the original HCS training) delivery across Portsmouth, delivered by author RAW. Table 1 provides a full description of the HCS ‘lite’ training according to the template for intervention description and replication (TIDieR) checklist (34). HCS training is based on social cognitive theory (35, 36), aiming to build trainees’ self-efficacy and confidence to have MECC conversations, and experiential learning, focusing on learning by practising and experiencing HCS (37). Subsequently, HCS training (including HCS ‘lite’) includes practise, modelling of the skills by the trainer, opportunities for reflection, and supportive group learning (37). The training content focusing on teaching the HCS philosophy (that is framed around motivating individuals to identify their own solutions) and the core healthy conversation skills to be applied to conversations. These four skills are; active listening (listening more than you speak), asking open discovery questions (ODQs) (open questions that spark motivation in that person and encourage them to come up with their own solutions), reflecting on practice, and facilitating SMARTER goal setting (helping people to set goals that are Specific, Measurable, Achievable, Relevant, and Time-bound, that are evaluated and reviewed at a later date) (23). The underlying message of the philosophy is to encourage and empower individuals to support their own wellbeing through positive behaviour change, as opposed to passively supporting them (e.g. through merely listening and providing empathy) or providing advice (21). HCS ‘lite’ training is focused on creating a supportive and non-judgemental setting. Although HCS ‘lite’ (a single training session) is pragmatically favoured to optimise the number of individuals who are able to attend, this means that there is no opportunity to implement HCS between sessions and therefore reflect on any alterations made to one’s practice within the training, which is concerning given that reflection is an important pillar of HCS training (37).

**Table 1:** Description (and programme architecture) of HCS ‘lite’ training organised using the template for intervention description and replication (TIDieR) checklist.

| **TIDieR checklist item** | **Description** |
| --- | --- |
| Name of the intervention | Healthy Conversation Skills 'lite' training |
| Why | Based on social cognitive theory and experiential learning pedagogy. The goal is to provide trainees with the confidence to deliver healthy conversations, by immersing themselves in the skills and approach. |
|  | • The philosophy is focused on four main pillars; 1) I am not responsible for the choices people make, 2) Being given information alone does not make people change, 3) People come to us with solutions, and 4) It is not possible to persuade people to change their habits.<br>• The four target skills are; asking open discovery questions (ODQs), active listening, supporting SMARTER goal setting, and reflection on practice. |
| What | **Procedure:**<br>• The training begins by centring itself around the unifying motivation to help people. The concept of MECC and its benefits are introduced briefly. The philosophy and skills are introduced using interactive activities.<br>• Throughout the training, trainees are encouraged to think holistically about their own wellbeing and what they could do to support it. E.g. trainees practise having a healthy conversation in pairs before and after the training content, and are encouraged to audio-record the conversations to listen back and reflect on them as a pair.<br>• Scenarios are presented and trainees are asked to respond by text on their phone and self-assess their response in accordance with the HCS approach. To facilitate psychological safety, HCS 'lite' deliberately does not include group feedback or modelling of an incorrect approach to a conversation.<br>• Trainees map influences of health onto the Dalgren and Whitehead social determinants of health model (38) during a group activity.<br>• Trainees are encouraged to consider one thing they will implement after the training.<br>**Materials:** A SMARTER goal setting sheet is provided. After training, trainees are provided with a cue card containing the philosophy and key skills. |
| Who Provided | Mostly a Public Health Practitioner from Portsmouth City Council. Some sessions delivered to undergraduate pharmacy students adopt a 'train the trainer' approach whereby previous cohorts of undergraduate pharmacy students were trained by RAW to deliver HCS 'lite' training to their peers. |
| How | In person, group-based (to allow for peer learning and support), usually in groups of around 6-30 people. Chairs are formed in an arc so that all trainees face each other, to facilitate discussion, avoiding use of notes and laptops, and aiding a supportive and non-judgmental group setting. |
| Where | Where all trainees are from the same organisation, training is delivered within a collaborative room within that organisation. For mixed groups (e.g. from some VCSE service providers), a room is booked that is accessible to all trainees (e.g. in a council building). |
| When and How Much | One three-hour session. |
| Tailoring | 'Train the trainer' sessions are sometimes tailored so that undergraduate pharmacy students can take ownership of the training. |
| Modifications | Due to timetabling, undergraduate pharmacy students generally receive HCS 'lite' training in large groups, whilst VCSE service providers often attend HCS 'lite' training as part of a mixed group containing service providers from a range of professions, although the training is also often delivered to specific VCSE organisations. |

Table 1 in Nichol et al. (19), which depicts regional MECC training according to the TIDieR checklist, illustrates the contrast with HCS training. Whilst both provide information about MECC and discuss health inequalities, regional MECC training focuses on providing information that can enable trainees to provide advice, whilst HCS (including HCS ‘lite’) training focuses on the skills needed to encourage individuals to identify their own needs for behaviour change.

### Development of initial programme theories (IPTs)

The development of IPTs for this realist evaluation was an iterative process, via four stages; 1) initial programme theory development based on retroduction and abductive reasoning (39), 2) refinement through informal literature scoping, 3) refinement through stakeholder involvement, and 4) prioritising IPTs.

Initial programme theory development began with drafting an overall theory of change for HCS ‘lite’ training, using Miro (40) to aid a mind mapping exercise. Initial ideas were organised into C, M, and O, and some initial tentative CM links were formed. This exercise began at the bottom level of abstraction (e.g. considering specific users) and informed more generalisable statements and proposed CM links, although at this stage no CMO configurations were complete as specific outcomes had not yet been configured. Mind maps were used to facilitate the grouping of contexts, mechanisms, and outcomes into more abstract and generalisable statements. This process also organised rival theories about when HCS ‘lite’ might not work, again organised by C, M, and O. This process also generated overarching inferences about how HCS ‘lite’ training works differently for different groups.

Next, scoping of the available literature on all iterations of HCS was conducted. Insights from the literature were systematically recorded into a table, organised by; study design, who (the sample), what (characteristics of HCS training), What worked (plus any thoughts about why), What did not work (plus any thoughts about why), and suggestions for programme theories (insights to inform the IPTs). Miro was again used to further develop higher order statements and start to link C, M, and Os. The statements were copied into a word document to facilitate linking of dyads (CM, CO, MO) and triads of CMO configurations. To further facilitate this process and help to illustrate how CMO configurations may interact with one another, a visual overall programme theory was created and then split and developed for each participant group specifically (VCSE service providers and undergraduate pharmacy students). Finally, a table was created to present CMO configurations alongside rival CMO configurations that illustrated how the outcomes could be different when the context was different, due to a differential triggering of mechanisms (see Supplementary Material 2). Throughout this process, a list of potentially relevant substantive theories (substantive theories (mid-range theories from specific domains that explain how and why mechanisms operate) was created, which will be revisited throughout data analysis. Use of existing substantive theories within realist evaluation facilitates the process of developing CMO configurations and enables knowledge translation by contributing to the evidence base to inform other programmes, beyond HCS training (41).

Stakeholder involvement further refined the IPTs. The initial CMO configurations and overall visual programme theories were shared with the deliverer of HCS ‘lite’ training (author RAW) and were refined iteratively until both authors agreed (BN and RAW). These were also shared with the study team (all authors) which further provided suggestions for refinement. Formal stakeholder involvement consisted of meetings with two groups of stakeholders; one with key decision-makers involved in the implementation of HCS (both original HCS and HCS lite) in the South East of England (henceforth referred to as stakeholders), and one with those who had received HCS ‘lite’ training (henceforth referred to as service providers). Stakeholders’ (n = 4; 3 female, 1 male) involvement in HCS covered its design, delivery, and implementation, including specifically within the VCSE. Due to attrition, all service providers (n = 3; all female) were undergraduate pharmacy students. Initial involvement of both groups highlighted two crucial areas of focus; 1) engaging individuals to attend HCS ‘lite’ training, and 2) adopting the HCS approach long-term. The initial IPTs were further edited, removed, and added to following stakeholder involvement. For example, an initially proposed mechanism was that during the ‘lite’ training, it is essential to be able to plan implementation of the HCS, and anything that is not planned will not be implemented. The feedback from both groups was that behavioural cues (e.g. discussions with colleagues, HCS posters, emails about HCS) after the training to prompt memory and associate current practice with the training was more important than forming intentions and planning during the training. Stakeholder involvement also helped to identify key differences in CMO configurations based on the participant group. For example, whilst the programme theories initially focused on comparing trainees with healthcare backgrounds (undergraduate pharmacy students) versus non-healthcare background (service providers from the VCSE), stakeholder involvement highlighted that a comparison was between trainees who have lots of experience with talking to service users (i.e. VCSE service providers) versus those who had much less experience (undergraduate pharmacy students).

Finally, a pragmatic approach to prioritising IPTs was taken (42), selecting IPTs according to priority areas identified through stakeholder involvement (engagement of trainees to attend HCS ‘lite’, and implementing HCS in practice long-term), ‘weak spots’ or areas where the desired outcome is not always produced, areas judged to show potential for the greatest public health impact (e.g. through facilitating behaviour change at scale), and holistically capturing outcomes of HCS ‘lite’ (from attending training to long-term change). Most importantly, IPTs were prioritised according to their relevance outside of the specific programme (HCS ‘Lite’ training) (43). Prioritisation resulted in 13 IPTs which were thematically grouped into five categories that describe different stages of HCS ‘lite’ training: 1) engagement (attending), 2) engagement (attention), 3) acquisition during training (pre-requisites to MECC delivery e.g. competence, confidence, and motivation), 4) applying the skills (delivering MECC), and 5) habit formation. The overall visual programme theory is shown in Figure 1 (see Supplementary Material 1 for the overall visual programme theories for each participant group), and the IPTs are shown in Supplementary Material 2. The overall IPT states that:

> *An individual engages with HCS because they are supported to by their organisation and the training is framed around their personal motives. HCS ‘lite’ training helps people to deliver health behaviour change conversations because trainees exchange their existing communication style with a new proactive and motivational approach, through reflection and immersion in the skills. However, to maintain the HCS approach after ‘lite’ training, trainees must be prompted through environmental cues and reminders, which is particularly impactful if trainees have already planned to deliver HCS during the training. Facilitation of SMARTER goal setting is not maintained because it does not align with an opportunistic approach, and so most trainees are ill equipped to implement this skill within their roles*.

**Figure 1:**
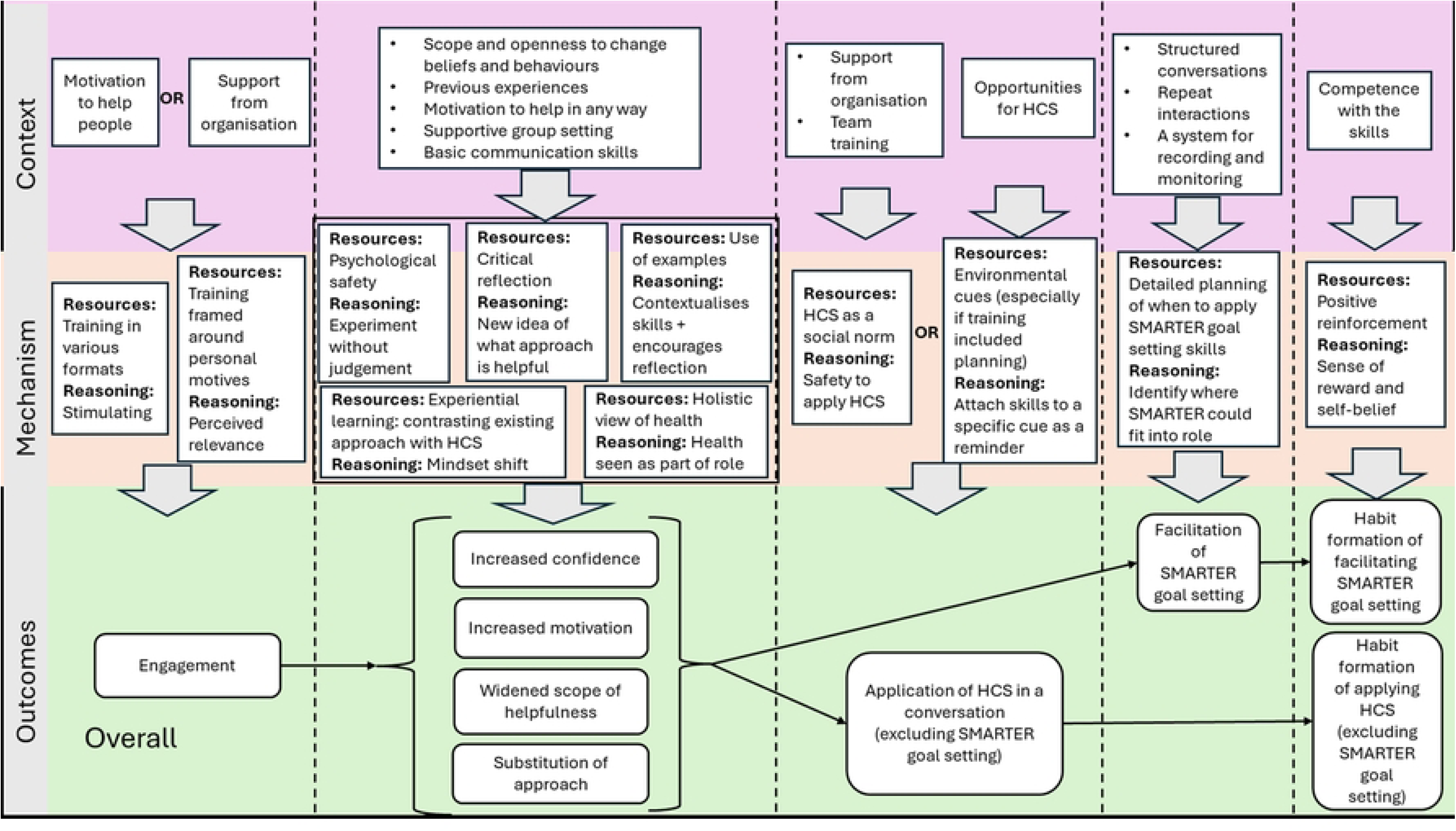
Visual depiction of the overall initial programme theory, for testing.

These propositions are exploratory, intended to be refined through iterative qualitative analysis and supported by quantitative data, consistent with the realist methodology.

### Quantitative pre- and post- survey

The quantitative before and after survey will be administered pre (immediately before receipt of training), post (immediately following training) and follow-up (8-10 weeks after training) to both participant groups. Opportunity sampling will be employed, such that all eligible trainees within the study period (August 2025 to April 2026) will be invited to complete the surveys. Where possible, the trainer (RAW) or coordinator of the training (e.g. a key contact at the University) will send the pre survey to trainees around 1 week before training. A QR code to complete the pre survey will also be displayed on the first slide of the training, with time provided at the start of the training for trainees to complete it. At the end of the training, a QR code to complete the post-survey will be provided and all trainees will be asked to complete it. Trainees will be encouraged to complete the post survey before leaving the training site, or at least within the next 24 hours following training (although will be able to access the link at any time). Eight weeks after the training, the trainer or coordinator of the training will send trainees a link to complete the follow-up survey. Since training to VCSE service providers is often as part of mixed professional groups and to avoid isolation of VCSE service providers, all attendees of mixed groups containing VCSE service providers will be invited to complete the surveys. Using Gpower to calculate the total sample size required to detect a significant difference in confidence, perceived usefulness, perceived importance, and competence between trainee groups, to detect a small effect size (0.3) at 80% power, a minimum total sample size of 352 (176 in each group) is required to detect a significant difference in confidence and competence post training between participant groups (two groups t test (correlations not available to calculate for multiple linear regression), two tailed, p < .05). With an added 10% margin to account for missing data (that is not missing at random), the target sample size is 387 (194 per group). If the target sample size is not reached, the available data will be analysed acknowledging that the analyses was underpowered as a limitation. As secondary analyses, differences pre and post within each group will also be tested.

Details of the survey items are available in the OSF pre-registration (https://osf.io/wpbn3/files). The consent form is embedded in the pre-survey and participants who do not consent will be automatically redirected to the end of the survey. Participants will be reminded at the start of the post- and follow-up survey that their participation indicates their continued consent. To link each participant’s responses to the three survey timepoints, participants will be instructed to create a structured code to re-enter at each survey timepoint. Aside from demographic information (e.g. age, gender, experience) which will be collected within the pre survey, the questionnaire is identical across the three timepoints. Perceived usefulness of MECC, perceived importance of MECC, confidence to deliver MECC conversations, and competence of using ODQs will be assessed using existing measurement tools employed in other evaluations of HCS training (21, 23–26). Additionally, the existing competence tool (26) has been adapted for the current study to also assess competence in facilitating SMARTER goal setting. Both measures of competence present four scenarios (e.g. "*I’ve lost count of the number of times I’ve tried to stop smoking—it’s hopeless*!") and ask participants to respond (in writing) as if they were having a real conversation with a client. After stakeholder involvement, a question was also included in the post-training survey timepoints to assess memory of the HCS philosophy. Each survey will take between 15-20 minutes to complete. At the end of the follow-up survey, survey participants will be invited to a) sign up to enter into a prize draw to win a £100 (first place), £50 (second place) or £25 (third place) Love2Shop voucher and b) complete an expression of interest form to take part in an interview.

Quantitative data will be analysed using SPSS (44). To create a mean score for competency of ODQs and SMARTER goal facilitation, responses to the four scenarios for each skill will be aggregately coded by two independent coders (BN and another author), using established coding matrices adapted from previous studies (26). Any discrepancies will be resolved through discussion with a third independent reviewer (AMR). Thus, aggregate scores (mean score across the four scenarios) will be calculated for both skills. Recall of the HCS philosophy will also be scored by two independent coders (BN and another author) and resolved through discussion with a third reviewer (AMR), using a self-created rubric available in the OSF pre-registration: https://osf.io/wpbn3/files. The data from attendees who are not undergraduate pharmacy students or service providers from the VCSE will not be included in the inferential analyses but will be summarised descriptively. Descriptive statistics will be produced for demographic characteristics, and pre and post scores relating to confidence, perceived usefulness, perceived importance, and competence (for both VCSE service providers and undergraduate pharmacy students). Exploratory baseline differences between professional groups for confidence, perceived usefulness, perceived importance, and competence will be tested through independent samples t-tests (or Mann-Whitney U tests if assumptions for an independent samples test are not met, tested using the Shapiro-Wilk test and Levene’s test for homogeneity of variances). Exploratory differences between trainee groups in post HCS ‘lite’ training (post survey) responses for perceived usefulness, perceived importance, and competence will be tested using a multiple logistic regression, controlling for baseline values (pre survey), age, and gender. Changes in confidence, perceived usefulness, perceived importance, and competence pre and post HCS ‘lite’ training will be tested within groups (conducted for both VCSE service providers and undergraduate pharmacy students) through within-subjects t-tests (or Wilcoxon Signed Rank tests If normality assumptions are not met, tested using the using Shapiro-Wilk test). Statistical testing will aid comparison with the existing literature of how HCS impact the confidence and competence of trainees across different settings. For the current realist evaluation, statistical testing will look to test (refine, support, refute) the IPTs, and therefore analysis will look to ‘preserve’ the configurations in analysis. For example, comparison across participant groups (undergraduate pharmacy students and VCSE service providers) will be tested for a relationship with proposed outcomes (motivation, competence, and confidence). If quantitative variables do not meet normality assumptions, non-parametric tests will be applied.

### Qualitative realist interviews

Qualitative semi-structured and theory driven realist interviews will be conducted with undergraduate pharmacy students and VCSE service providers who have received receipt of HCS ‘lite’ training. In accordance with a realist approach (45), purposive sampling will be applied to sample participants from a broad range of contexts identified as relevant from the IPTs (e.g. types of role within the VCSE, interval since receipt of HCS ‘lite’ training). The purposive sampling approach will also be iterative depending on relevant contexts that emerge throughout the interviews. Participants will be recruited via two sources; those who complete the expression of interest form after the follow-up survey, and through contacting previously trained pharmacy students and VCSE service providers. In accordance with a realist approach which focuses on explanatory depthbased on coherence and credible causality, defining the sample size a priori for each participant group was not possible (45, 46). However, to assist with funding acquisition for the project, information power (47) was applied to provide an estimate of the target sample size for each participant group. The aim is specific and the lead researcher (BN) is an experienced qualitative interviewer, although this must be balanced with the possibility of convenience sampling depending on interest and that the analysis will test programme theories, which would both require a larger sample size. Thus, the estimated sample size is 40 (n = 20 per professional group), although the exact final sample size and characteristics of the sample will be developed iteratively.

Topic guides have been structured around the IPTs, seeking to test and refine them whilst also being open to emergent programme theories. As is distinctive with realist interviews, interviews (guided by the topic guide) will adopt a teacher learning cycle, whereby the researcher (BN) shares their initial theorising (most often parts of their IPTs in lay terms) and asks the participant to share their thoughts (46). Participants become the teacher by sharing their own thoughts out how HCS training worked for them (46). One topic guide has been developed for each participant group (VCSE service providers and undergraduate pharmacy students), given that the overall programme theories differ between groups. Broadly, both topic guides cover the context in which participants work, how they engaged with the training, experienced the training, and applied and embedded the training. Again, in accordance with a realist approach the content of the topic guide will be iterative, to reflect the evolving programme theories. Both initial topic guides are available on the OSF pre-registration (https://osf.io/wpbn3/files).

Participants will be sent the consent form for interviews and asked to return written consent prior to the interview. Interviews will be offered online via Teams but may also be conducted in person if needed. At the start of the interview, consent will be confirmed verbally. Interviews will likely last between 30-60 minutes and will be audio recorded (if in-person) or recorded in-app (if on Teams). Participants that take part in an interview will be renumerated with a £20 shopping voucher for approximately one hour of their time. Recordings will be sent to a transcription service for transcription, then anonymised by the lead author (BN). In accordance with open science practices, anonymised transcripts will be made publicly available via the UK Data Service, providing participants provide their consent.

The data will be analysed iteratively to test, refine, and add to the IPTs using CMO configurations. Transcripts will firstly be imported into NVivo Version 12 (48) for analysis. The planned data analysis approach within Nvivo has been adapted from existing guidance (49), and may be further amended once data analysis begins. Data coding will be organised by IPTs, through creating a node for each IPT. Emerging programme theories will be added with new nodes. Analysis will preserve dyads and triads of CMO configurations to ensure a coherent analysis. Codes to organise data within each node (programme theory) will be developed iteratively, but will distinguish whether the data supports of refutes it. Subsequently, the writeup will distinguish between unsubstantiated (there was no relevant data relating to that IPT) and discounted (there was data refuting that IPT) programme theories. Mind mapping (e.g. using Miro) and editing of the existing overall programme theories will be utilised to develop causal understanding throughout analysis. Analysis will be conducted by the lead author (BN) although updates to programme theories will be discussed with the research team and stakeholders (46, 50). Changes to the IPTs throughout the analysis will be documented using linked Memos, using highlighting for new additions and strikethroughs for deletions. Again, substantive/formal theory will be utilised to assist with explanatory understanding. Theories will be sourced through the existing list created during IPT development, conversations with experts (all authors), and further literature searching. Inferences will be drawn from the depth and richness of the data in informing programme theories and the coherence and credible causality provided by the programme theories themselves.

Following existing guidance for realist evaluations (46), several quality assurances will be adopted. Given that realist interviewing should be informed by naturalistic observation, the lead author observed HCS ‘lite’ training which informed the initial development of IPTs. Although repeat interviews will not be feasible given the existing requirement of repeated surveys, the lead author (BN) will instead encourage participants to email if they develop any further theories after the interview.

### Triangulation

The survey will not specifically test IPTs but will provide validation of the proposed outcomes, organised broadly by context of trainees (comparing VCSE service providers and undergraduate pharmacy students). The qualitative data will instead link the outcomes identified within the survey to context and mechanisms. Both streams of data will be triangulated through ‘qualitising’ (transforming into qualitative data) the quantitative survey data (51). Namely, C, M, or O (most likely) statements of configurations will be generated, generating dyads of CMO configurations where possible (e.g. that a specific context leads to a specific outcome). These will be added as a file into Nvivo and coded in the same way as the transcripts (as provided above), so that the quantitative data can be fully incorporated into the qualitative analysis.

### Optimising involvement, inclusion, and engagement

Realist methods are grounded in a pragmatic aim of policy change (52). Thus, accessing individuals with relevant insights and translating the findings into policy ad practice is arguably even more central and important than for other methods. Optimising inclusion of individuals with relevant insights was considered. HCS ‘lite’ training is delivered in the English language across Portsmouth, so the evaluation assumes a fundamental understanding of the English language of participants. Thus, translation services will not be available. Paper copies of the survey will be available to trainees during HCS ‘lite’ training. Dissemination of the study findings will aim to utilise a range of formats for accessibility, including an animation. Stakeholder involvement will further inform appropriate dissemination strategies.

### Knowledge translation

As this study was co-designed with author RAW, the findings of this evaluation will directly inform the approach to HCS adopted across Portsmouth. Additionally, involvement of key decision makers has created ownership of the project which will facilitate impact. Authors BN, AMR, and CH are also established researchers of MECC subsequently have developed connections with other implementors of MECC nationally. These existing networks will be utilised to disseminate the findings to other regions, to help them consider their approach to MECC training. Furthermore, the findings of the current evaluation aim to inform a future effectiveness study of MECC delivery, through providing HCS training. It was first necessary to ensure that HCS enables all trainees to deliver MECC conversations.

## Discussion

This realist evaluation will produce a refined programme theory which explains how and why a specific approach to MECC training, HCS, works, including how it works differently for different audiences and thus how it might need to be adapted to ensure that all trainees are equipped equally to deliver MECC conversations. By including both undergraduate pharmacy students and VCSE service providers, this study will enable comparison based on a number of potentially important contextual factors. VCSE service providers do not work in a healthcare setting and may not have a healthcare background or any prior knowledge of health promotion, so may not view MECC as relevant to them (28). However, they often have highly diverse specialist backgrounds and are experienced and confident in talking to service users, strongly driven by a desire for social good (28). Meanwhile, undergraduate pharmacy students are trained as healthcare professionals, including to deliver healthy lifestyle advice, with health promotion now considered a key part of a Pharmacist’s role (53). However, undergraduate pharmacy students may have had little experience of interacting with service users and have chosen to study pharmacy for a range of reasons (54). The realist evaluation allows for these contextual differences to be incorporated into the analysis, to provide further understanding about what elements of HCS work for whom and why. The findings will be published in a peer reviewed journal, submitted prior to April 2027. Additionally, the findings will be disseminated with key stakeholders (including the stakeholder involvement group for this study), providing recommendations to optimise impact of the evaluation.

The main anticipated challenge for the study is achieving the target sample size for the quantitative surveys, to reach sufficient statistical power to detect for significance. However, differences pre and post HCS ‘lite’ training can still be tested for within-groups with a smaller sample, which can be narratively compared between participant groups. Equally, the main strength of this study is that it has been co-produced between research and practice, making knowledge translation more likely. This context has maintained a pragmatic focus for the current evaluation. For example, visual depictions of programme theories are essential to communicate the findings with individuals outside of research settings. The challenge of co-production approaches is maintaining the collaboration (55), which has been facilitated within this project by in-person research visits. One limitation is that given that the study was not specifically designed to test programme theories, the quantitative data will not statistically test CMO configurations. Whilst the two participant groups will allow for significant differences in outcomes between groups of trainees to be tested, the IPTs highlight that the context is far more nuanced than occupation or job role. The qualitative data will thus be essential in configuring the outcomes identified in the survey with their respective context and mechanisms.

Nevertheless, the key target outcome of the current realist evaluation is to inform approaches to MECC training across the UK and beyond. Ensuring that MECC training is effective in helping people to deliver MECC conversations may help to create a culture around health promotion and prevention. There is currently no specific guidance about how MECC training is delivered, indicating the potential for policy-level impact of the current study. For example, if HCS is demonstrated to increase confidence and competence no matter the background of trainees, there is rationale for MECC policy to specify HCS training as the approach to MECC delivery. Alternatively, both approaches may instead complement each other, providing a rationale to include elements of both. MECC is also unique in that it raises the possibility of health promotion outside of healthcare settings, opening new opportunities to prevent disease. Due to trusting relationships within the VCSE, there is rationale that MECC within the VCSE may be more impactful than when delivered in other settings (e.g. healthcare) where the service provider is not known to the service user. Assessing whether HCS ‘lite’ training is sufficient to enable service providers from the VCSE to deliver MECC is the first step to evaluating the impact of MECC within the VCSE. Also, by utilising existing measures to assess the change in confidence, motivation, and competence following HCS ‘lite’ training, the results of the survey will contribute to the existing literature supporting HCS ‘lite’ training, and aid comparison between other trainee groups (e.g. staff at Sure Start children’s centres, physiotherapists) (21, 29). Thus, the current study will provide a holistic understanding of what elements of HCS training work for whom, under what circumstances, in what respects, to what extent, and why, and in the process informing the same questions about MECC training overall.

## Data Availability

No datasets were generated or analysed during the current study. All relevant data from this study will be made available upon study completion.

